# Application of Machine Learning Approaches to Develop Predictive Models for Diabetes and Hypertension among Bangladesh Adults

**DOI:** 10.1101/2025.05.30.25328660

**Authors:** Gulam Muhammed Al Kibria, James O’Hagan, Golam Shariar, Tarina Khan, Mohammed Elfaramawi

## Abstract

**Introduction:** With rapid urbanization, lifestyle changes, and an aging population, non-communicable diseases (NCDs), including hypertension and diabetes, pose significant public health challenges in Bangladesh and many other low- and middle-income countries. This study used machine learning (ML) approaches to develop predictive models for hypertension and diabetes among adults in this country.

**Methods:** We analyzed Bangladesh Demographic and Health Survey 2022 data. This is a nationally representative cross-sectional survey. Participants were classified as hypertensive when their systolic blood pressure was ≥140 mmHg, diastolic blood pressure was ≥90 mmHg, or if they used antihypertensive medication. They were classified as diabetic if their fasting plasma glucose was ≥7.0 mmol/L or they used glucose-lowering drugs. Potential predictors included age, gender, education, wealth quintile, overweight/obesity, rural-urban residence, and division of residence. Descriptive analysis was conducted, and six ML models were applied: artificial neural network (ANN), random forest, adaptive boosting (AdaBoost), gradient boosting, XGBoost, and support vector machine (SVM). Models’ performance was evaluated via accuracy, area under the curve (AUC), sensitivity, specificity, and F1-score. Feature importance was assessed to rank risk factors.

**Results:** The study included 13,847 adults, 55% of whom were females. Diabetes and hypertension had prevalence rates of 16.3% and 20.5%, respectively, with both conditions increasing with age, and the highest prevalence was 24.4% for diabetes and 43.3% for hypertension in individuals aged 65 and older. Wealthier and urban residents experienced higher rates (diabetes: 24.9% among the richest compared to 9.9% among the poorest; hypertension: 23.3% in urban versus 19.2% in rural areas). Additionally, overweight/obesity was a strong predictor for both conditions. For diabetes, AdaBoost had the highest AUC (0.699) and SVM had the highest accuracy (0.836); for hypertension, AdaBoost had the greatest AUC (0.775) and accuracy (0.799). Hypertension topped diabetes predictors, while overweight/obesity led for hypertension, followed by age and diabetes. Wealth and gender were moderately influential, with education and geographic factors less so. Low specificity across models indicated challenges in identifying non-cases.

**Conclusion:** In this ML-driven analysis, we identified the bidirectional relationship of hypertension and diabetes along with several other predictors, including overweight/obesity, older age, and richer household wealth quintiles. Our findings underscore the need for integrated screening and lifestyle interventions targeting high-risk groups to mitigate future NCD burden.

## Introduction

Non-communicable diseases (NCDs) such as hypertension and diabetes have emerged as critical public health challenges globally, with a particularly alarming rise in low- and middle-income countries (LMICs), including Bangladesh [1–3]. Recent estimates suggest that NCDs are responsible for over two-thirds of global deaths each year, with cardiovascular diseases and diabetes ranking among the top contributors [4,5]. Rapid urbanization, sedentary lifestyles, poor dietary habits, and an aging population have caused the increasing prevalence of these conditions in LMICs, including Bangladesh [1,6,7]. According to the International Diabetes Federation (IDF), approximately 13.1 million adults in Bangladesh were living with diabetes in 2021, a number projected to grow significantly by 2045 if current trends persist [8]. Similarly, hypertension affects nearly one in four adults in the country, often undiagnosed until complications arise [9,10]. These results underscore the urgent need for proactive strategies to identify at-risk populations and implement effective prevention measures.

Artificial intelligence (AI) and machine learning (ML) have revolutionized the analysis of complex health data, and offer a data-driven approach to identify patterns and predictors of diseases or outcomes which are not identifiable by conventional statistical methods [11,12]. For instance, unlike traditional regression models, ML techniques such as decision trees, random forests, and neural networks can handle high-dimensional data and detect non-linear relationships [11].

ML has been successfully applied in various settings to predict burden for conditions like diabetes and hypertension based on factors like body mass index (BMI), family history, and lifestyle habits [13,14]. The lack of data on conditions like hypertension and diabetes was a problem in Bangladesh and other similar LMICs. However, multiple recent surveys and studies have investigated the burden and factors affecting these two conditions [3,6,15,16]. For instance, studies reported that people with older age and higher socioeconomic status (i.e., higher education and wealth) have a higher burden of hypertension/diabetes than those with younger age and lower socioeconomic status [6,16,17]. Despite the growing body of research on NCDs in Bangladesh, several gaps in the literature persist, including the identification of actionable risk factors through advanced analytics like ML [18,19]. This narrow scope overlooks the diversity of risk factors across rural and urban populations, as well as socioeconomic gradients captured in national surveys like the BDHS. Although global studies have highlighted predictors such as obesity and wealth status using ML techniques, there is a lack of evidence on how these factors differ in a Bangladeshi setting. These gaps highlight the need for a comprehensive, ML-driven analysis that uses nationally representative data to provide a holistic understanding of hypertension and diabetes risk.

To overcome these limitations, this study employs ML techniques on the BDHS 2022 dataset. Our objectives are twofold: (1) to identify predictors of diabetes and hypertension among Bangladeshi adults, and (2) to assess the performance of different ML models in this area. We aim to reveal new insights into the factors driving these conditions and inform focused public health strategies. The results have the potential to help policymakers develop interventions that tackle the underlying causes of NCDs, thereby easing their impact on Bangladesh’s healthcare system.

## Methods

### Study design

We analyzed data from BDHS 2022, a nationwide cross-sectional survey in Bangladesh. This survey covered rural and urban regions of all administrative divisions of Bangladesh. It aimed to obtain estimates of major demographic and health indicators, including hypertension and diabetes. Mitra and Associates, a private research firm in Bangladesh, conducted it. Approximately two hundred data collectors underwent recruitment and training to conduct interviews. Data collection took place from June to December of 2022 [9]. We analyzed the data for the present study in April 2025.

### Sample design and coverage

BDHS 2022 used a multistage cluster sampling design. It created a sampling frame based on Bangladesh’s 2011 housing and population census. This sample frame contained a list of enumeration areas (EA). Households were chosen from the EA in two stages, and data were collected during household visits [9].

For BDHS 2022, 675 EAs were chosen, comprising 425 from rural and 250 from urban areas [10,20]. A higher number of EAs were selected from rural regions to align with the country’s population distribution. Approximately 30 households were randomly selected from each EA in the second stage. One-third of the households were eligible for blood pressure and blood sugar measurements. These ensured the representativeness of the samples to ensure reliable estimates of demographic and health indicators. The age eligibility for BDHS 2022 hypertension and diabetes modules was 18. The response rate for the survey was 98%. Detailed information on both BDHS, including survey designs, methodologies, sample size estimations, questionnaires, and results, can be found online [9].

### Outcome variables

The outcome variables of the present study were hypertension and diabetes.

### Hypertension

We considered a person hypertensive if s/he met at least one of these three characteristics: systolic blood pressure (SBP) of 140 mmHg or above, diastolic blood pressure (DBP) of 90 mmHg or above, and the participant was getting any hypertensive medication. LIFE SOURCE® UA-767 Plus monitors recorded the blood pressure (BP). Three measurements were done with ten minutes of interval between each measurement. Other data were collected during the interval. The mean of the last two BP measurements was used to mark the final BP [9].

### Diabetes

We considered a person diabetic if s/he had a fasting plasma glucose of 7.0 mmol/l (or above) or was taking blood sugar-lowering medications. The HemoCue 201+ blood analyzer was used to record the glucose [9].

### Potential Factors

Based on scientific plausibility, literature search, and data structure, we investigated the following variables as potential features associated with the outcomes: women’s age, education level, household wealth quintile, overweight/obesity, and rural-urban place and division of residence. Since diabetes and hypertension have a bidirectional association (i.e., they can increase the risk of one another) [21–23], we investigated hypertension as a risk factor for diabetes and diabetes as a risk factor for hypertension.

Age was grouped as 18-34, 35–44, 45–54, 55-64, and 65 or more years. The education level was classified as no formal education, primary (i.e., 1 to 5 school years), secondary (i.e., 6 to 10 school years), and college/above education (i.e., 11 or more school years). Respondents reported their current work status. The household wealth index score was obtained by principal component analyses of households’ basic building materials (e.g., floors, roofs, and walls), drinking water sources, sanitation facilities, and availability of electricity and other items. The score was then stratified into quintiles: poorest, poorer, middle, richer, and richest. Dhaka, Chittagong, Rajshahi, Khulna, Barisal, Rangpur, Sylhet and Mymensingh were the eight divisions during BDHS 2022 [9].

### Statistical Analysis

All analyses were performed using Python. First, we conducted descriptive analyses and calculated the distribution (%) of diabetes and hypertension across the features listed above. We used chi-square tests to examine the association of these factors with hypertension and diabetes. We divided the dataset into training (80%) and test sets (20%). As the dataset was imbalanced (i.e., low prevalence of diabetes/hypertension), we applied synthetic minority over-sampling technique (SMOTE) and adaptive synthetic sampling (ADASYN) to balance the training dataset. SMOTE generates synthetic samples for the minority class by interpolating between existing minority samples [24], and ADASYN does the same but focuses more on harder-to-learn minority samples near decision boundaries [25]. We implemented six ML algorithms for classification: (1) Artificial Neural Network (ANN); (2) Random forest; (3) AdaBoost; (4) Stochastic Gradient Boosting (GBM); (5) Extreme Gradient Boosting (XGBoost); and (6) Support Vector Machine (SVM).

We used 10-fold cross-validation during training and validation to check model reliability. We used preprocessing to transform categorical variables via one-hot encoding. Model hyperparameters were fine-tuned through a combination of grid search and cross-validation

We calculated accuracy, sensitivity, specificity, area under the curve (AUC), and F1-score to assess and compare the performance of the six ML models. We also used receiver operating characteristic (ROC) curves to evaluate the discriminative abilities of the models.

For algorithms capable of estimating feature importance (e.g., random forest, XGBoost, GBM), we identified the most significant predictors using feature importance scores. For linear models like SVM (with a linear kernel), we extracted model coefficients to determine each variable’s impact on hypertension and diabetes prediction. Bar plots were used to visually represent the coefficients and feature importance rankings, improving interpretability.

Data manipulation tasks (e.g., recoding or grouping) were handled using the pandas and numpy libraries. Visualizations were created with Matplotlib and Seaborn, while the ML algorithms were developed using scikit-learn, XGBoost, and TensorFlow/Keras libraries.

## Results

A total of 13,835 adults were included in the analysis; about 16.3% and 20.6% had diabetes and hypertension, respectively (Table 1). Diabetes affected 15.6% of males and 16.9% of females (P=0.052). The burden of hypertension was significantly higher in females (23.5%) compared to their male counterparts (17.0%; P<0.001). Prevalence of both conditions increased with age (P<0.001), peaking at 24.4% for diabetes (65+ years) and 43.3% for hypertension (65+ years). Education showed a modest association with diabetes (P=0.046), while hypertension was highest among those with no education (28.6%; P<0.001). Wealth quintiles revealed a gradient, with diabetes rising from 9.9% among the poorest to 24.9% among the richest and hypertension from 15.4% (poorest) to 27.6% (richest) (P<0.001). Urban residents had higher rates (diabetes: 21.7%; hypertension: 23.3%) than rural residents (13.5%; 19.2%; P<0.001). Divisionally, Dhaka had the highest diabetes prevalence (21.1%), while Rajshahi had the highest prevalence for hypertension (23.6%; P<0.001). Overweight/obesity significantly increased both conditions’ prevalence (P<0.001).

**Table 1:**
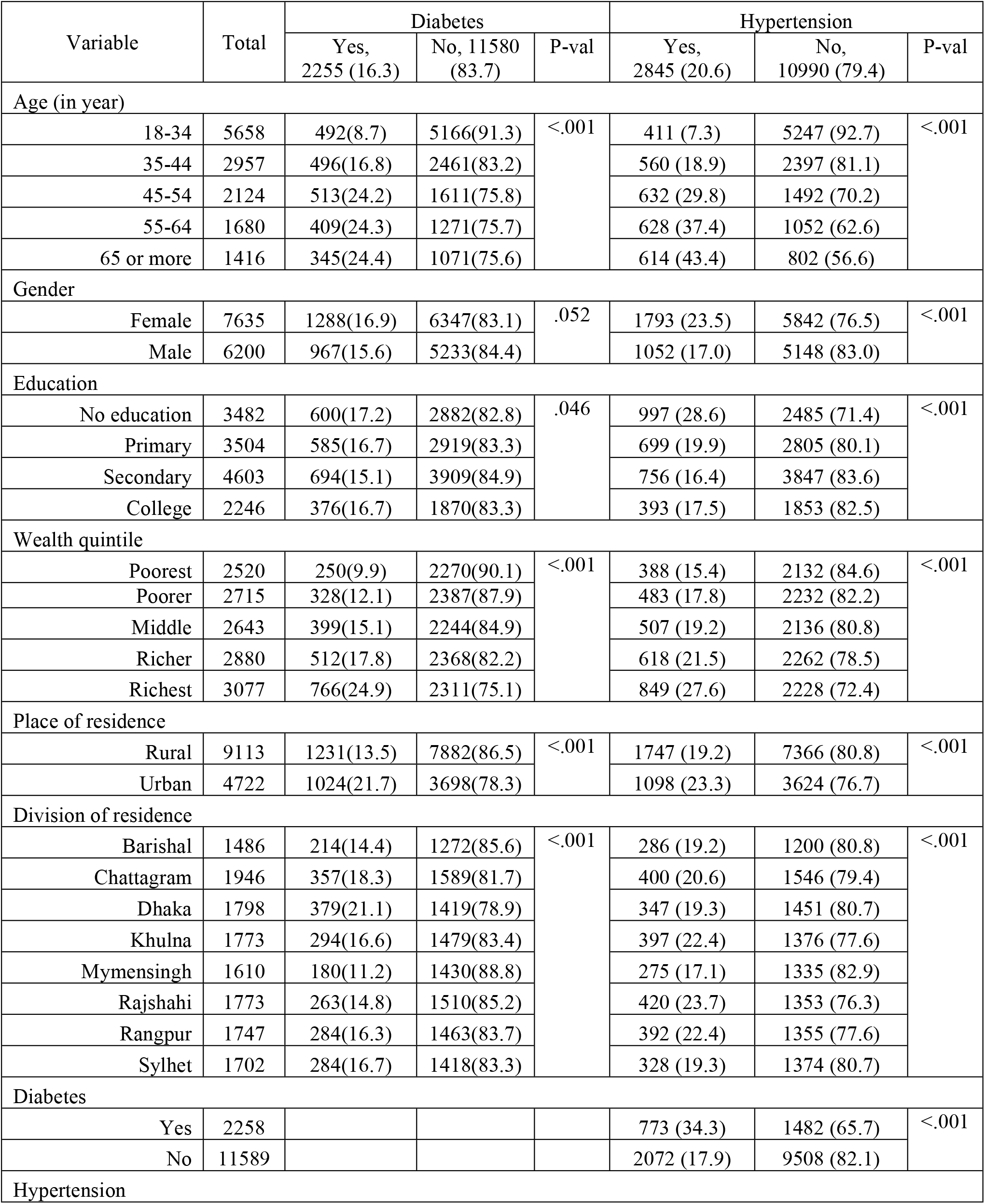
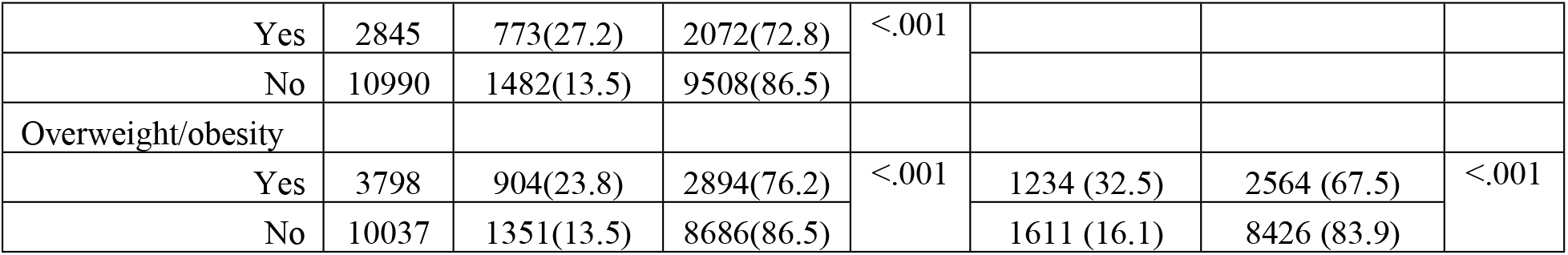
Distribution of diabetes and hypertension according to features.

Table 2 shows the performance metrics of six ML models. For diabetes, XGBoost achieved the highest accuracy (0.821) and F1 score (0.899), with a sensitivity of 0.958, indicating strong positive class detection, though specificity was low (0.127). Gradient boosting followed closely (accuracy: 0.803, F1: 0.885). SVM and random forest also performed well, with accuracies of 0.805 and 0.790, respectively. AdaBoost had the lowest accuracy (0.682) but a higher AUC (0.694). For hypertension, XGBoost again outperformed others (accuracy: 0.779, F1: 0.865), with a sensitivity of 0.899. Gradient boosting and SVM showed balanced performance (accuracies: 0.751, 0.755). AdaBoost had the highest AUC (0.770) but lower accuracy (0.700). Overall, XGBoost consistently demonstrated superior predictive performance across both conditions, particularly in sensitivity and F1 score.

**Table 2:** Performance metrices of the evaluation models obtained by ADASYN.

| Outcome | Model | Accuracy | AUC | Precision | Sensitivity | Specificity | F1 Score |
| --- | --- | --- | --- | --- | --- | --- | --- |
| Diabetes | ANN | 0.759 | 0.585 | 0.857 | 0.854 | 0.284 | 0.856 |
|  | Random Forest | 0.790 | 0.636 | 0.848 | 0.911 | 0.179 | 0.879 |
|  | ADA Boost | 0.682 | 0.694 | **0.891** | 0.706 | **0.563** | 0.787 |
|  | Gradient Boosting | 0.803 | **0.695** | 0.863 | 0.909 | 0.271 | 0.885 |
|  | XGBoost | **0.821** | 0.652 | 0.847 | **0.958** | 0.127 | **0.899** |
|  | SVM | 0.805 | 0.632 | 0.859 | 0.916 | 0.245 | 0.887 |
| Hypertension | ANN | 0.683 | 0.680 | 0.850 | 0.726 | 0.526 | 0.783 |
|  | Random Forest | 0.750 | 0.704 | 0.830 | 0.858 | 0.349 | 0.844 |
|  | ADA Boost | 0.700 | 0.770 | **0.899** | 0.698 | **0.711** | 0.786 |
|  | Gradient Boosting | 0.751 | **0.773** | 0.876 | 0.797 | 0.582 | 0.835 |
|  | XGBoost | **0.779** | 0.745 | 0.833 | **0.899** | 0.333 | **0.865** |
|  | SVM | 0.755 | 0.737 | 0.862 | 0.820 | 0.514 | 0.840 |

In Figure 1, we presented ROC curves to evaluate the discriminative ability of six classification models. For diabetes, gradient boosting achieved the highest AUC (0.695), followed closely by AdaBoost (0.694), indicating strong overall performance in distinguishing classes. XGBoost (AUC: 0.652) and SVM (0.632) showed moderate performance, while ANN had the lowest AUC (0.585). Random forest (AUC: 0.636) performed slightly better than ANN but lagged behind the top models. For hypertension, Gradient Boosting again led with the highest AUC (0.773), closely followed by AdaBoost (0.770) and XGBoost (0.745), reflecting robust classification capabilities. SVM (AUC: 0.737) and random forest (0.704) showed moderate performance, while ANN had the lowest AUC (0.680). Across both conditions, Gradient boosting consistently demonstrated superior discriminative power, with AdaBoost also performing notably well, particularly in AUC metrics.

**Figure 1.**
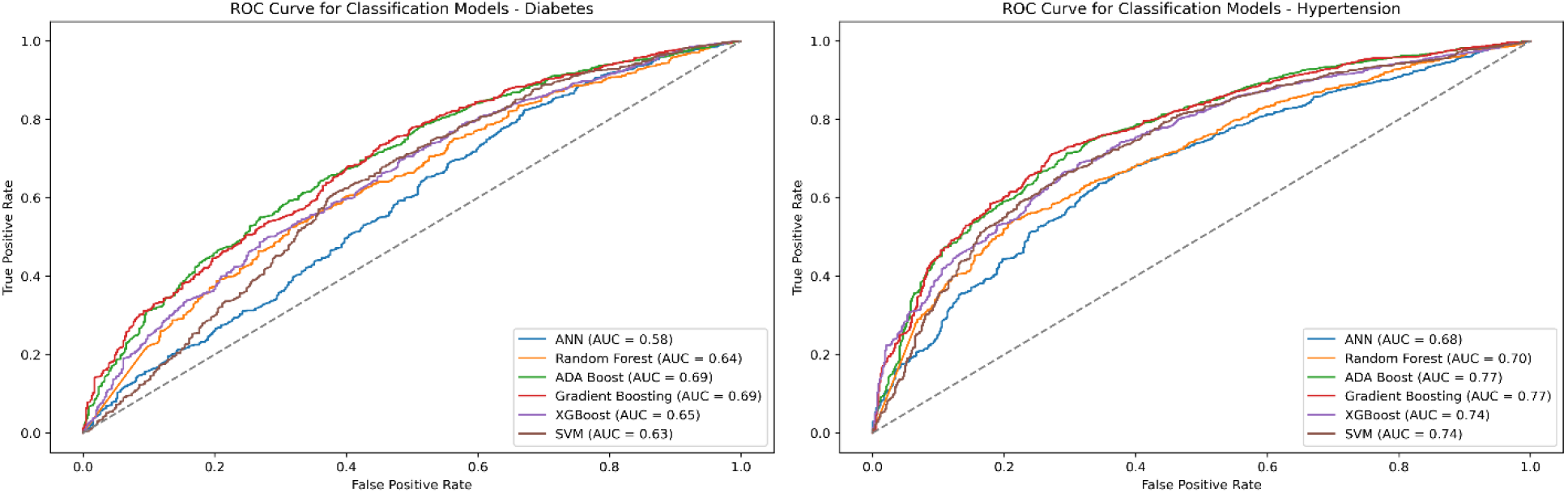
Receiver operating characteristics curve for the outcomes.

We reported feature importance for predicting diabetes an Table 3. We reported the features that were among the top 5 for 6 models. Notably, the top 5 features for both outcome variables were similar. For diabetes, hypertension ranked first across all models (frequency=6), followed by overweight/obesity (second, frequency=6) and age (third, frequency=6). Wealth quintile and gender ranked fourth–fifth, while education, division, and place of residence ranked sixth–eighth. For hypertension, overweight/obesity was the top predictor (first, frequency=6), followed by age (second, frequency=6) and diabetes (third, frequency=6). Wealth quintile and gender ranked fourth–fifth, with education, division, and place of residence at sixth–eighth. Health-related features (hypertension, overweight/obesity, and diabetes), and age consistently dominated. Education and geographic factors showed lower importance, with SVM slightly varying in ranking wealth quintile and gender. These findings highlight key risk factors for diabetes and hypertension, informing targeted interventions.

**Table 3:** The top 5 identified features in all studied models.

| Features | Frequency<br>as top 5<br>features<br>among 6<br>models | Rank in Particular Model |  |  |  |  |  |
| --- | --- | --- | --- | --- | --- | --- | --- |
|  |  | ANN | Random<br>Forest | AdaBoost | Stochastic<br>Gradient<br>Boosting | XGBoost | SVM |
| Diabetes |  |  |  |  |  |  |  |
| Hypertension | 6 | 1 | 1 | 1 | 1 | 1 | 1 |
| Overweight/<br>Obesity | 6 | 2 | 2 | 2 | 2 | 2 | 2 |
| Age | 6 | 3 | 3 | 3 | 3 | 3 | 3 |
| Wealth Quintile | 6 | 4 | 4 | 4 | 4 | 4 | 5 |
| Gender | 6 | 5 | 5 | 5 | 5 | 5 | 4 |
| Education | 0 | 6 | 7 | 6 | 7 | 7 | 6 |
| Division | 0 | 7 | 6 | 7 | 6 | 6 | 7 |
| Place of residence | 0 | 8 | 8 | 8 | 8 | 8 | 8 |
| **Hypertension** |  |  |  |  |  |  |  |
| Overweight/obesity | 6 | 1 | 1 | 1 | 1 | 1 | 1 |
| Age | 6 | 2 | 2 | 2 | 2 | 2 | 2 |
| Diabetes | 6 | 3 | 3 | 3 | 3 | 3 | 3 |
| Wealth Quintile | 6 | 4 | 4 | 4 | 4 | 4 | 5 |
| Gender | 6 | 5 | 5 | 5 | 5 | 5 | 4 |
| Education | 0 | 6 | 7 | 6 | 7 | 7 | 6 |
| Division | 0 | 7 | 6 | 7 | 6 | 6 | 7 |
| Place of residence | 0 | 8 | 8 | 8 | 8 | 8 | 8 |

## Discussion

We used nationally representative data from BDHS 2022 and applied modern ML approaches to identify and rank predictors of diabetes and hypertension among Bangladeshi adults. In addition to observing the bidirectional relationship of diabetes and hypertension, we found that age, wealth quintile, and overweight/obesity are potential associated features for the studied conditions in Bangladesh. Our findings not only confirmed known risk factors for diabetes or hypertension but also provided insights into the demographic, socioeconomic, and health-related variables that impact these two outcomes in this country. These insights have important implications for targeted prevention, policy development, and health system planning.

From a methodological perspective, the use of ML algorithms allowed for robust predictive modeling. The identification of complex relationships among variables might have been overlooked using traditional statistical techniques. AdaBoost, XGBoost, and gradient boosting consistently outperformed other models in terms of sensitivity and F1 scores for both conditions. Their superior performance underscores the value of ensemble learning methods in health prediction tasks, specifically with multidimensional data [11,12]. SVM and ANN also performed well, though SVM exhibited the lowest AUC for diabetes, indicating weaker discriminative ability for that outcome. Despite the application of SMOTE and ADASYN, the low specificity observed across models, particularly for diabetes, highlights a limitation in distinguishing non-cases, could result from low prevalce of both outcomes (i.e., 16% for diabetes and 21% hypertension). This low specificity levels suggest that further model refinement and perhaps inclusion of additional behavioral or clinical variables may enhance performance [11,12].

The feature importance rankings revealed a consistent pattern across all ML models. Health-related variables (i.e., hypertension, diabetes, and overweight/obesity) and age were top predictors. Wealth quintile and gender also had significant association, while education, place of residence, and division had relatively lesser influence [6,16,26]. This hierarchy of predictors reinforces the notion that while demographic and geographic contexts matter, physiological and lifestyle-related indicators are the most immediate determinants of NCD risk [16,27].

The prevalence estimates by features observed in this study underscore the significance of known predictors that could be used to develop and implement prevention and control programs in the context of this country. For instance, consistent with prior research, both conditions showed a strong association with older age, and individuals aged 65 years and above have the highest burden [6,26,28]. This age-associated rise is consistent with biological aging processes and cumulative exposure to risk factors such as obesity, poor diet, and sedentary behavior [18]. Although women had a higher burden of hypertension, the diabetes burden was not significantly higher. These gendered differences may reflect differences in healthcare access, behavioral risk profiles, and sociocultural dynamics [29,30].

Among the most important predictors across all six ML models were hypertension for diabetes (and vice versa). This finding reaffirmed the bidirectional and interdependent relationship between diabetes and hypertension [21,22]. Previous studies have documented that hypertension increases insulin resistance, and diabetes can impair vascular function, which ultimately increases blood pressure levels [21,22,30]. Our findings support integrating screening and management protocols for both conditions simultaneously, specifically in primary and outpatient care settings in upazila (i.e., sub-district) health complexes, district hospitals, or medical college hospitals.

Overweight/obesity (i.e., BMI 25kg/m^2^ or above) appeared as a predictor for both diabetes and hypertension across all models. This echoed global evidence on the central role of excess adiposity in metabolic dysfunction [31,32]. Cardiometabolic syndromes can also occur due to adipocyte hypertrophy, visceral adiposity, and ectopic fat deposition. Additionally, compensatory insulin secretion is closely associated with obesity [33]. The transition from pre-diabetes to type 2 diabetes is caused by this hypersecretion, which also causes insulin resistance and ultimately β-cell failure [32,33]. The overall prevalence of overweight/obesity has increased in Bangladesh. For instance, the prevalence of overweight/obesity was 25% among 35+-year-old people in 2011, this increased to 32% in 2022 [9,20]. The rising prevalence has been driven by urbanization, dietary transitions, and physical inactivity and poses a significant challenge to the ongoing burden of NCDs like diabetes and hypertension [1,17]. Public health campaigns promoting healthy lifestyles, including physical activity and nutritional education, are urgently needed to curb this trend.

Wealth quintile, a proxy for socioeconomic status, demonstrated a “dose-response relationship” with both conditions; disease prevalence increased from the poorest to the richest households. Another predictor variable, education level, often considered as a socioeconomic variable in addition to wealth quintile, did not have a significant association with either of the conditions. This is contrary to previous research and suggests that the significance of education level on the burden of hypertension/diabetes is declining [16,26]. The wealth pattern likely reflects a shift in the burden of diabetes/hypertension toward wealthier segments of the population due to their greater access to energy-dense diets, sedentary occupations, and urban environments. However, this does not diminish the vulnerability of populations with lower household wealth, who may face more barriers to receiving early diagnosis and effective treatment [34]. Socioeconomic gradients in NCD prevalence call for a dual approach to prevent or control these conditions. It signifies targeting high-risk behaviors in affluent groups while ensuring equitable access to care for the poor.

Our study fills critical gaps in the Bangladeshi literature on NCDs. Unlike earlier studies that primarily focused on prevalence estimates, we employed predictive analytics to identify actionable risk factors. Additionally, by incorporating ML techniques into nationally representative data analysis, this study moves beyond traditional epidemiological approaches, offering a more nuanced and dynamic understanding of disease risk. This methodology also allows for potential future integration into health decision-support systems, enabling personalized risk assessments and real-time public health surveillance.

Limitations of this study also merit discussion. First, the cross-sectional design of BDHS restricts causal inference. Second, some important behavioral and clinical predictors (e.g., dietary intake, physical activity levels, and family history) were not available in the dataset, potentially limiting model accuracy. Third, the models were trained on structured categorical variables; incorporating continuous or unstructured data in future analyses may yield improved performance. Lastly, although ML offers powerful tools for prediction, the black-box nature of some algorithms (e.g., ANN) may hinder interpretability for clinical application.

## Conclusion

This study demonstrates that survey data with advanced ML models can effectively identify key predictors of hypertension and diabetes. Our results highlight the dominant role of modifiable risk factors such as overweight/obesity and wealth, in addition to the reinforcing relationship between the two conditions. These findings should guide the design of targeted, data-driven public health interventions that prioritize high-risk groups, promote early screening, and support healthy lifestyle changes to reduce the burden of NCDs across Bangladish.

## Data Availability

Data is available from the DHS website: https://dhsprogram.com/data/available-datasets.cfm

## Declarations

### Abbreviations

ADASYN: nAdaptive synthetic sampling
BMI: Body mass index
BP: Blood pressure
CKD: Chronic kidney disease
GFR: Glomerular filtration rate
HDL: High density lipoprotein
LDA: Linear discriminant analysis
ML: Machine learning
NHANES: National Health and Nutrition Examination Survey
SMOTE: Synthetic Minority Over-sampling Technique
SVM: Support vector machine.

## Acknowledgment

The authors thank the survey participants for their time.

## Consent for publication

Not applicable

## Availability of data and material

Data is available from https://dhsprogram.com/data/available-datasets.cfm

## Ethics, Consent to Participate, and Consent to Publish declarations

Not applicable.

## Authors contributions

Conceptualization: Gulam Muhammed Al Kibria

Data Cleaning and Analysis: Gulam Muhammed Al Kibria

Investigation: Gulam Muhammed Al Kibria, Golam Shariar, Tarina Khan, James O’Hagan, Mohammed Elfarawami

Methodology: Gulam Muhammed Al Kibria, James O’Hagan

Writing-Original Draft: Gulam Muhammed Al Kibria

Writing – Review & Editing: Golam Shariar, Tarina Khan, James O’Hagan, Mohammed Elfarawami, Mohammed Elfarawami

